# The Phenotypic Spectrum of *COL4A3* Heterozygotes

**DOI:** 10.1101/2023.04.11.23288298

**Authors:** Kaushal V. Solanki, Yirui Hu, Bryn S. Moore, Vida Abedi, Venkatesh Avula, Tooraj Mirshahi, Regeneron Genetics Center, Natasha T. Strande, Ion D. Bucaloiu, Alexander R. Chang

## Abstract

Most data on Alport Syndrome (AS) due to COL4A3 are limited to families with autosomal recessive AS or severe manifestations such as focal segmental glomerulosclerosis (FSGS). Using data from 174,418 participants in the Geisinger MyCode/DiscovEHR study, an unselected health system-based cohort with whole exome sequencing, we identified 403 participants (0.2%) who were heterozygous for likely pathogenic COL4A3 variants. Phenotypic data was evaluated using International Classification of Diseases (ICD) codes, laboratory data, and chart review. To evaluate the phenotypic spectrum of genetically-determined autosomal dominant AS, we matched COL4A3 heterozygotes 1:5 to non-heterozygotes using propensity scores by demographics, hypertension, diabetes, and nephrolithiasis. COL4A3 heterozygotes were at significantly increased risks of hematuria, decreased estimated glomerular filtration rate (eGFR), albuminuria, and end-stage kidney disease (ESKD) (p<0.05 for all comparisons) but not bilateral sensorineural hearing loss (p=0.9). Phenotypic severity tended to be more severe among patients with glycine missense variants located within the collagenous domain. For example, patients with Gly695Arg (n=161) had markedly increased risk of dipstick hematuria (OR 9.47, 95% CI: 6.30, 14.22) and ESKD diagnosis (OR 7.01, 95% CI: 3.48, 14.12) whereas those with PTVs (n=119) had moderately increased risks of dipstick hematuria (OR 1.63, 95% CI: 1.03, 2.58) and ESKD diagnosis (OR 3.43, 95% CI: 1.28, 9.19). Less than a third of patients had albuminuria screening completed, and fewer than 1/3 were taking inhibitors of the renin-angiotensin-aldosterone system (RAASi). Future studies are needed to evaluate the impact of earlier diagnosis, appropriate evaluation, and treatment of ADAS.

**Significance Statement:** Alport Syndrome (AS) is the second most common genetic cause of end-stage kidney disease (ESKD), yet little is known about the penetrance and phenotypic spectrum of genetically-determined Autosomal Dominant AS. Using an unselected health system-based cohort, we compared individuals heterozygous for likely pathogenic or pathogenic variants in COL4A3 to a propensity score-matched control group and demonstrate increased risks of hematuria, albuminuria, and ESKD. Risks of kidney disease phenotypes were markedly elevated for missense glycine variants in the collagenous domain and moderately elevated for those with PTVs, compared to controls. The vast majority had not been diagnosed with AS and less than a third ever received albuminuria testing, suggesting opportunities to improve management by early genetic diagnosis.

## Introduction/Background

Alport Syndrome (AS), the second most common cause of monogenic kidney failure, is characterized by hematuria, sensorineural deafness and ocular abnormalities. Historically, about 85% of AS cases have been due to X-linked COL4A5, with the remainder mostly due to autosomal recessive disease from COL4A3 and COL4A4 variants.^1,2^ AS develops because the COL4A3, COL4A4 and COL4A5 genes encode the alpha-3, alpha-4 and alpha-5 subunits of type IV collagen—the major structural component of glomerular basement membrane (GBM). Heterozygous carriers for autosomal recessive and X-linked conditions are often asymptomatic. However, recent data have shown that heterozygous carriers for COL4A3, COL4A4, and COL4A5 (females only) can present with a wide spectrum of renal manifestations, ranging from familial microscopic hematuria, thin basement membrane nephropathy (TBMN), to more severe manifestations such as focal segmental glomerulosclerosis (FSGS) and end-stage kidney disease (ESKD).^3^ Up to 40% of patients with TBMN and 10% of FSGS cases have pathogenic or likely pathogenic heterozygous variants in COL4A3, COL4A4 or COL4A5.^4,5^ As most studies include cohorts enriched with the most severely affected individuals, the prevalence, penetrance and full phenotypic spectrum of AS remain unknown.

Advances in molecular genetics have led to the use of genetic testing in confirming the diagnosis of AS, and expert opinions suggest that genetic testing for COL4A3, COL4A4, and COL4A5 variants could take precedence over renal biopsy when AS is suspected or if there is significant family history of AS-related kidney disease.^6^ Genetic testing with next generation sequencing gene panels is also recommended in patients with steroid-resistant FSGS.^7^ What remains less clear is the role of earlier genetic diagnosis of autosomal dominant Alport Syndrome (ADAS), especially as costs of exome sequencing decrease and availability of testing increases. More information is needed on individuals with undiagnosed disease who could potentially receive early treatment with angiotensin converting enzyme inhibitors (ACEi) to reduce future risk of ESKD.^8 9,10^

In this study, we examine the penetrance and phenotypic spectrum of heterozygous COL4A3 pathogenic variants using data from GeisingerMyCode-DiscovEHR study, an unselected health system-based cohort. We hypothesized that heterozygous carriers of COL4A3 pathogenic variants would be at increased risks of hematuria, albuminuria, FSGS and ESKD.

## Methods

### Study Population

The Geisinger Institutional Review Board approved this study. Informed consent was waived as participants were previously consented in the MyCode™ Community Health Initiative as part of the Geisinger-Regeneron DiscovEHR collaboration. ^11^ For this study we included 174,361 participants in MyCode who had whole exome sequencing data available, and a COL4A3 variant that was classified as pathogenic (P) or likely pathogenic (LP) previously in ClinVar.^12^

### Whole Exome Sequencing (WES) and Variant calling

WES was performed in collaboration with Regeneron Genetics Center, as previously described.^13^ A modified version of the xGEN probe from Integrated DNA Technologies (IDT) were used for target sequence capture (**Supplemental Methods**). ^14^ We included COL4A3 variants that were listed in ClinVar at least once as P/LP (accessed 5/25/2021), with any number of ClinVar stars. To examine potential genotype-phenotype comparisons, we categorized P/LP variants into 4 separate categories: 1) Gly695Arg, which was the most common P/LP variant; 2) other glycine missense variants located within the collagenous domain (between the end of exon 2 and the beginning of exon 48)^15^ since glycine variants are critical in the intermediate collagenous domain; 3) protein truncating variants (PTVs); 4) other missense variants or inframe deletions. Varsome (www.varsome.com accessed 2/23/23), a human genomic variant search engine that provides automated classification of pathogenicity using American College of Medical Genetics (ACMG) criteria was used to report predicted pathogenicity. We also documented whether COL4A3 P/LP heterozygotes had any additional rare (allele frequency <0.001) variants in COL4A3, COL4A4, or COL4A5.

### Phenotyping

Electronic health record (EHR) data including demographic characteristics, International Classification of Diseases (ICD) 9 and 10 diagnosis codes and laboratory data were extracted. We linked data to the United States Renal Data System (USRDS) to ascertain end-stage kidney disease (ESKD) status and presumed cause of ESKD,^16^ and also used a definition including ICD diagnosis codes for dialysis or kidney transplant. Due to USRDS data restrictions, we report only data using ESKD per ICD code for subgroups <11. ICD diagnosis codes used for ESKD, hematuria, bilateral sensorineural hearing loss, and FSGS are listed in **Supplemental Table 1**. Laboratory data included serum creatinine, urinalysis, urine albumin/creatinine ratio (ACR), and urine protein/creatinine ratio (PCR). To minimize false positive hematuria tests that could be due to urinary tract infection, we included only urinalyses negative for leukocyte esterase and nitrites. Urinalysis-based hematuria outcomes were categorized as trace or greater blood and 1+ or greater if present on ≥50% of urinalyses. Urinalysis-based proteinuria outcomes were categorized as 1+ or greater or 2+ or greater if on at least 2 urinalyses. Albuminuria was defined as having moderate albuminuria (ACR 30-299 mg/g or PCR 150-499 mg/g), or severe albuminuria (ACR 300+ mg/g or PCR 500+ mg/g). Estimated glomerular filtration rate (eGFR) was calculated using the CKD-EPI 2021 formula.^17^

To provide a comprehensive phenotypic data as available in the EHR, chart review was performed on the 402 COL4A3 carriers by KS with additional review of patients with kidney biopsies by AC and IDB. We searched for additional data on AS-related phenotypic features, family history of AS and TBMD, audiometry, treatment with angiotensin converting enzyme inhibitors (ACEis) or angiotensin receptor blockers (ARBs), kidney biopsies, urologic workup of hematuria, and genetic testing.

### Outcomes

The primary outcome was having any phenotypic feature of AS (hematuria on urinalysis or ICD code, dipstick proteinuria 1+, moderate albuminuria, severe albuminuria, eGFR <60 and <30 ml/min/1.73m^2^, FSGS, ESKD, or bilateral sensorineural hearing loss). Secondary outcomes included each of these outcomes separately, as well as Kidney Disease Improving Global Outcomes (KDIGO) risk categories,^18^ which ranged from no CKD, hematuria alone, moderately increased risk, high risk, very high risk, and extremely high risk. Only individuals with eGFR and urinalysis data were included in the KDIGO risk category analysis; if quantitative ACR was unavailable, we classified dipstick protein 1+ twice as ACR 30-299 mg/g, and dipstick protein 2+ or greater twice as ACR 300+ mg/g.

### Statistical Analysis

We first compared phenotypic features between COL4A3 P/LP heterozygotes and the rest of the MyCode cohort. As urinalysis testing was done more frequently in COL4A3 P/LP heterozygotes we performed propensity score matching to reduce potential selection bias since hematuria is a key feature of AS. We used a combination of propensity score based on age, Black race, Hispanic ethnicity, and exactly-matched categories (sex, hypertension, diabetes, nephrolithiasis, and year of first outpatient encounter) to match COL4A3 P/LP variant heterozygotes 1:5 to individuals without COL4A3 P/LP variants, using RStudio (Version 2022.02.3). The standardized mean difference (SMD) was calculated to measure the balance of covariates between COL4A3 P/LP heterozygotes and the matched control cohort. Categorical and continuous outcomes were compared between heterozygotes and non-heterozygotes using chi square and t-tests, respectively. Logistic regression was used to estimate the adjusted odds ratios for AS-related phenotypes using STATA/MP 15.1. We also examined whether outcomes differed between the 4 COL4A3 subgroups, using chi square tests. Logistic regression analyses comparing subgroups to the overall control group were adjusted for age, sex, and race. We used complete case analyses.

## Results

Out of 174,361 MyCode participants, 5993 (3.4%) had COL4A3 variants at minor allele frequency <1%, including 403 (0.2%) who had a COL4A3 variant previously reported in Clinvar as P/LP at least once. After excluding 1 COL4A3 P/LP heterozygote due to missing data, we examined 402 heterozygotes in detail (**Figure 1**). The most common P/LP COL4A3 variant was Gly695Arg (n=161). There were 47 patients with other glycine missense variants located within the collagenous domain, 119 with PTVs, and 75 with other missense variants or inframe deletions (n=73) (**Supplemental Table 2 for additional details by variant**).

**Figure 1.**
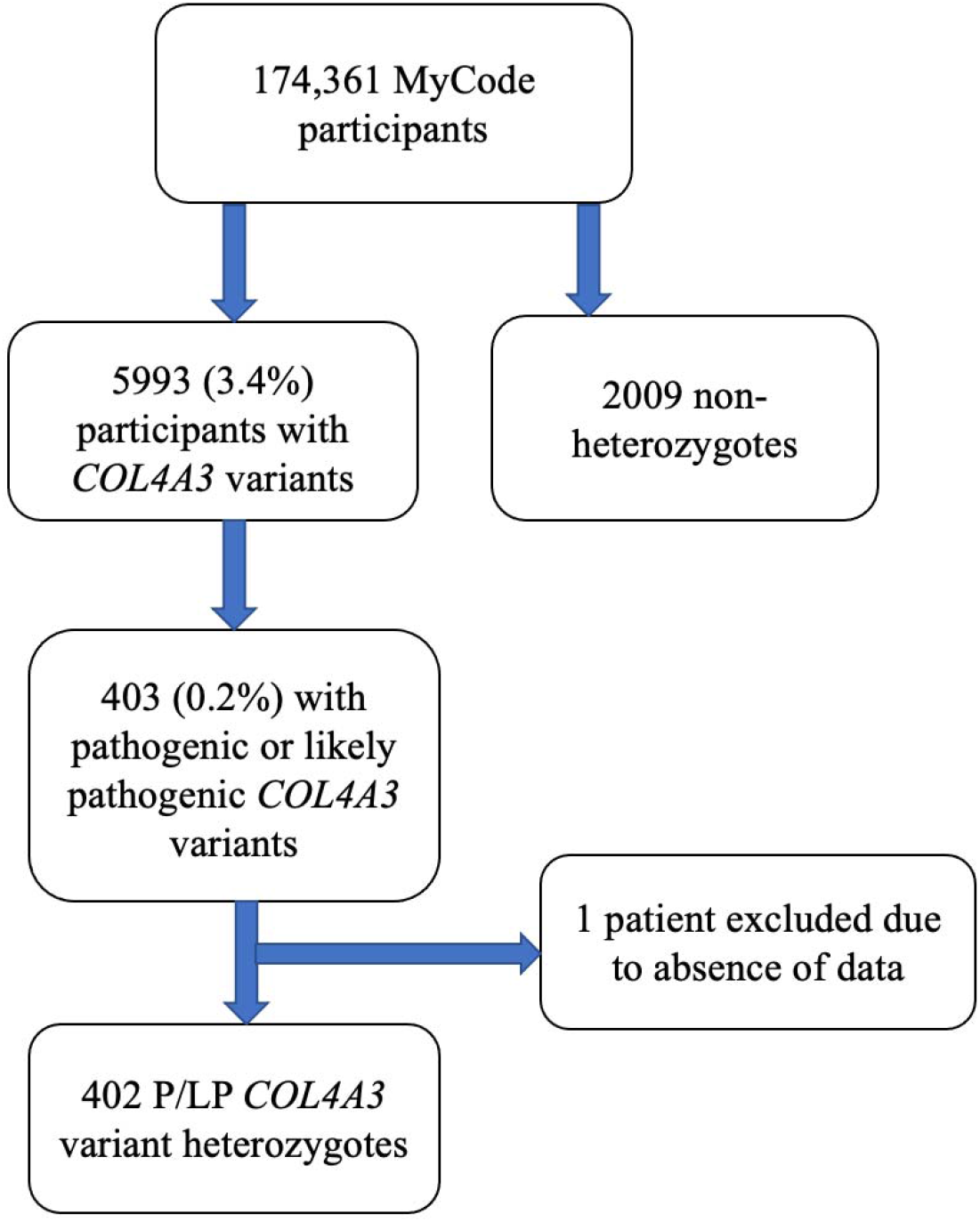
Flowchart. This figure does not use USRDS data.

Compared to the rest of the MyCode cohort, COL4A3 P/LP heterozygotes were more likely to have undergone urinalysis evaluation (81.1% vs. 75.7%; p=0.01) but had similar rates of evaluation for eGFR (89.1% vs. 88.8%; p=0.9) and ACR (29.4% vs. 28.1%; p=0.6), and similar prevalence of hypertension, diabetes, and nephrolithiasis (**Table 1**). After successfully matching 402 COL4A3 P/LP heterozygotes to 2009 non-heterozygote controls using a 1:5 ratio, mean age was 59.1 years and 64% were female, with similar urinalysis availability (81.1% vs. 77.7%; p=0.2), and median follow-up time (15 years, interquartile interval 7.8-18.6). Compared to controls, COL4A3 P/LP heterozygotes were more likely to have at least one phenotypic feature (64.4% vs. 45.7%; p<0.001) compared to controls with significant differences by variant group (chi-square p<0.001) (**Table 2, Supplemental Table 3**). All AS phenotypic features (hematuria, albuminuria, decreased eGFR, FSGS, and ESKD) were significantly higher in COL4A3 P/LP heterozygotes compared to controls (p<0.05 for all comparisons; **Supplemental Table 3**), except for bilateral sensorineural hearing loss.

**Table 1.**
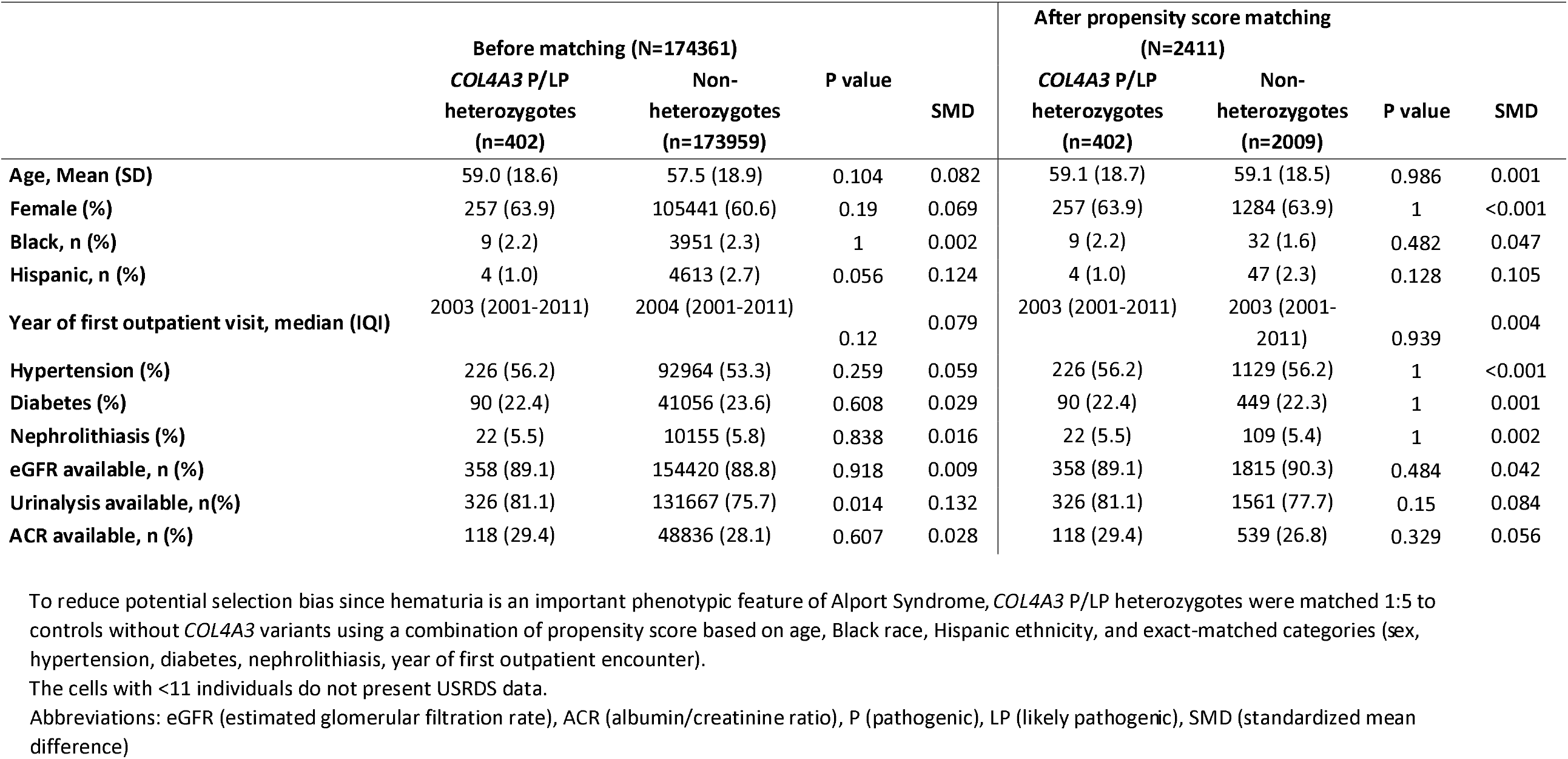
Characteristics of 402 COL4A3 P/LP heterozygotes and non-heterozygotes before and after matching.

**Table 2.**
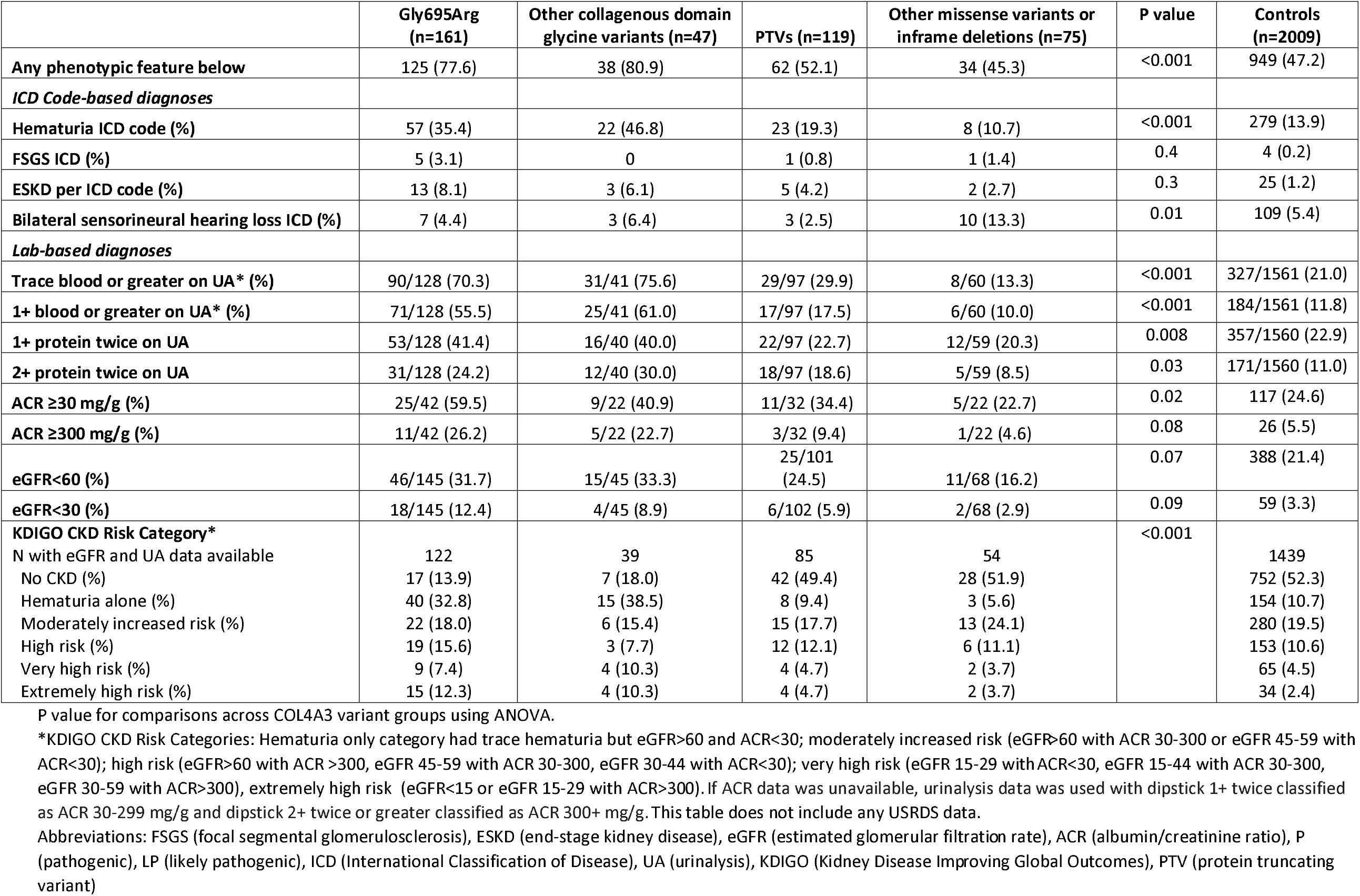
Phenotypic Features of COL4A3 P/LP Variant Groups and Controls.

|  | Gly695Arg<br>(n=161) | Other collagenous domain<br>glycine variants (n=47) | PTVs (n=119) | Other missense variants or<br>inframe deletions (n=75) | P value | Controls<br>(n=2009) |
| --- | --- | --- | --- | --- | --- | --- |
| **Any phenotypic feature below** | 125 (77.6) | 38 (80.9) | 62 (52.1) | 34 (45.3) | <0.001 | 949 (47.2) |
| *ICD Code-based diagnoses* |  |  |  |  |  |  |
| Hematuria ICD code (%) | 57 (35.4) | 22 (46.8) | 23 (19.3) | 8 (10.7) | <0.001 | 279 (13.9) |
| FSGS ICD (%) | 5 (3.1) | 0 | 1 (0.8) | 1 (1.4) | 0.4 | 4 (0.2) |
| ESKD per ICD code (%) | 13 (8.1) | 3 (6.1) | 5 (4.2) | 2 (2.7) | 0.3 | 25 (1.2) |
| Bilateral sensorineural hearing loss ICD (%) | 7 (4.4) | 3 (6.4) | 3 (2.5) | 10 (13.3) | 0.01 | 109 (5.4) |
| *Lab-based diagnoses* |  |  |  |  |  |  |
| Trace blood or greater on UA* (%) | 90/128 (70.3) | 31/41 (75.6) | 29/97 (29.9) | 8/60 (13.3) | <0.001 | 327/1561 (21.0) |
| 1+ blood or greater on UA* (%) | 71/128 (55.5) | 25/41 (61.0) | 17/97 (17.5) | 6/60 (10.0) | <0.001 | 184/1561 (11.8) |
| 1+ protein twice on UA | 53/128 (41.4) | 16/40 (40.0) | 22/97 (22.7) | 12/59 (20.3) | 0.008 | 357/1560 (22.9) |
| 2+ protein twice on UA | 31/128 (24.2) | 12/40 (30.0) | 18/97 (18.6) | 5/59 (8.5) | 0.03 | 171/1560 (11.0) |
| ACR ≥30 mg/g (%) | 25/42 (59.5) | 9/22 (40.9) | 11/32 (34.4) | 5/22 (22.7) | 0.02 | 117 (24.6) |
| ACR ≥300 mg/g (%) | 11/42 (26.2) | 5/22 (22.7) | 3/32 (9.4) | 1/22 (4.6) | 0.08 | 26 (5.5) |
| eGFR<60 (%) | 46/145 (31.7) | 15/45 (33.3) | 25/101<br>(24.5) | 11/68 (16.2) | 0.07 | 388 (21.4) |
| eGFR<30 (%) | 18/145 (12.4) | 4/45 (8.9) | 6/102 (5.9) | 2/68 (2.9) | 0.09 | 59 (3.3) |
| **KDIGO CKD Risk Category\*** |  |  |  |  | <0.001 |  |
| N with eGFR and UA data available | 122 | 39 | 85 | 54 |  | 1439 |
| No CKD (%) | 17 (13.9) | 7 (18.0) | 42 (49.4) | 28 (51.9) |  | 752 (52.3) |
| Hematuria alone (%) | 40 (32.8) | 15 (38.5) | 8 (9.4) | 3 (5.6) |  | 154 (10.7) |
| Moderately increased risk (%) | 22 (18.0) | 6 (15.4) | 15 (17.7) | 13 (24.1) |  | 280 (19.5) |
| High risk (%) | 19 (15.6) | 3 (7.7) | 12 (12.1) | 6 (11.1) |  | 153 (10.6) |
| Very high risk (%) | 9 (7.4) | 4 (10.3) | 4 (4.7) | 2 (3.7) |  | 65 (4.5) |
| Extremely high risk (%) | 15 (12.3) | 4 (10.3) | 4 (4.7) | 2 (3.7) |  | 34 (2.4) |

P value for comparisons across COL4A3 variant groups using ANOVA.
\*KDIGO CKD Risk Categories: Hematuria only category had trace hematuria but eGFR>60 and ACR<30; moderately increased risk (eGFR>60 with ACR 30-300 or eGFR 45-59 with ACR<30); high risk (eGFR>60 with ACR >300, eGFR 45-59 with ACR 30-300, eGFR 30-44 with ACR<30); very high risk (eGFR 15-29 with ACR<30, eGFR 15-44 with ACR 30-300, eGFR 30-59 with ACR>300), extremely high risk (eGFR<15 or eGFR 15-29 with ACR>300). If ACR data was unavailable, urinalysis data was used with dipstick 1+ twice classified as ACR 30-299 mg/g and dipstick 2+ twice or greater classified as ACR 300+ mg/g. This table does not include any USRDS data.
Abbreviations: FSGS (focal segmental glomerulosclerosis), ESKD (end-stage kidney disease), eGFR (estimated glomerular filtration rate), ACR (albumin/creatinine ratio), P (pathogenic), LP (likely pathogenic), ICD (International Classification of Disease), UA (urinalysis), KDIGO (Kidney Disease Improving Global Outcomes), PTV (protein truncating variant)

### Phenotypic Variability by P/LP COL4A3 Genotype

There was significant variability between the 4 genotype subgroups for having any phenotypic feature present as well as the number of phenotypic features present (**Tables 2,3**). Penetrance and phenotypic severity was highest in glycine missense variants located within the collagenous domain: Gly695Arg (n=161; 78% any phenotypic feature, 70% trace blood on dipstick, 32% eGFR <60, 12% eGFR <30, 3% FSGS, 8% ESKD per ICD code); other glycine collagenous domain variants (n=47; 81% any phenotypic feature, 76% trace blood on dipstick, 33% eGFR<60, 9% eGFR<30, 6% ESKD per ICD code).

Penetrance was lower in PTVs (n=119; 52% any phenotypic feature, 30% trace blood on dipstick, 25% eGFR<60, 6% eGFR<30, and 4% ESKD per ICD code). Results were similar among PTVs at earlier vs. later exons, including the most common PTV Gly1602AlafsTer13 (2/32 [6%] ESKD per ICD code) (**Supplemental Table 3**). Patients with other missense or inframe deletions (n=75) did not have significantly higher risk of most AS phenotypes (45% any phenotypic feature, 13% trace blood on dipstick, 16% eGFR <60, 3% eGFR<30, 3% ESKD per ICD code (**Tables 2,3**), other than bilateral sensorineural hearing loss (13%). There were 26 patients who had an additional heterozygous rare variant in COL4A3, COL4A4, or COL4A5 with 4 (15.4%) having ESKD ICD diagnosis (**Supplemental Table 4**).

**Table 3.** Risks of Alport Syndrome Phenotypic Features.

|  | **Gly695Arg** | **Other collagenous**<br>**domain glycine**<br>**variants** | **PTVs** | **Other missense**<br>**variants or**<br>**inframe deletions** |
| --- | --- | --- | --- | --- |
| **Any phenotypic feature below** | 4.45 (2.98, 6.63)*** | 5.18 (2.98, 6.63)*** | 1.17 (0.80, 1.73) | 0.99 (0.61, 1.61) |
| *Any ICD diagnoses* |  |  |  |  |
| **Hematuria ICD code** | 3.44 (2.43, 4.88)*** | 5.65 (3.12, 4.88)*** | 1.46 (0.91, 2.35) | 0.77 (0.36, 1.62) |
| **FSGS ICD** | 15.62 (4.13, 59.15)*** | - | 4.12 (0.45, 37.27) | 7.45 (0.81, 68.02) |
| **ESKD per ICD code** | 7.01 (3.48, 14.12)*** | 5.83 (1.67, 20.41)** | 3.43 (1.28, 9.19)* | 2.33 (0.54, 10.13) |
| **ESKD per USRDS** | 8.96 (4.21, 19.04)*** | 5.08 (1.13, 22.74)* | 2.85 (0.82, 9.88) | 3.14 (0.71, 13.86) |
| **Bilateral sensorineural hearing loss** | 0.77 (0.35, 1.69) | 1.23 (0.37, 4.05) | 0.43 (0.14, 1.40) | 2.91 (1.44, 5.91)** |
| *Lab-based diagnoses* |  |  |  |  |
| **Trace blood or greater on UA\*** | 9.47 (6.30, 14.22)*** | 12.55 (6.02, 26.13)*** | 1.63 (1.03, 2.58)* | 0.62 (0.29, 1.33) |
| **1+ blood or greater on UA\*** | 9.50 (6.46, 13.96)*** | 12.11 (6.31, 23.25)*** | 1.60 (0.93, 2.78) | 0.88 (0.37, 2.09) |
| **1+ protein twice on UA** | 2.21 (1.53, 3.18)*** | 2.40 (1.28, 4.50)** | 1.05 (0.65, 1.70) | 0.90 (0.47, 1.72) |
| **2+ protein twice on UA** | 2.41 (1.56, 3.71)*** | 3.85 (1.96, 7.59)*** | 1.99 (1.17, 3.37)* | 0.77 (0.30, 1.96) |
| **ACR ≥30 mg/g** | 4.74 (2.47, 9.10)*** | 2.25 (0.93, 5.44) | 1.66 (0.78, 3.54) | 0.89 (0.32, 2.47) |
| **ACR ≥300 mg/g** | 6.20 (2.81, 13.69)*** | 4.58 (1.56, 13.41)*** | 1.80 (0.51, 6.28) | 0.84 (0.11, 6.48) |
| **eGFR<60** | 1.88 (1.21, 2.93)** | 2.76 (1.31, 5.83)** | 1.09 (0.64, 1.85) | 0.75 (0.35, 1.58) |
| **eGFR<30** | 4.32 (2.41, 7.74)*** | 3.32 (1.10, 10.02)* | 1.76 (0.72, 4.26) | 0.96 (0.22, 4.10) |

Logistic regression analyses comparing each variant subgroup to control group, adjusted for age, sex, and race.
\*P<0.05; \*\*P<0.01; \*\*\*P<0.001
Abbreviations: FSGS (focal segmental glomerulosclerosis), ESKD (end-stage kidney disease), USRDS (United States Renal Data System), UA (urinalysis), eGFR (estimated glomerular filtration rate), ACR (albumin/creatinine ratio), ICD (International Classification of Disease), PTV (protein truncating variant)

Odds ratios (ORs) of having any phenotypic feature were increased for Gly695Arg (OR 4.45, 95% CI: 2.98, 6.63; p<0.001) and other collagenous domain glycine variant groups (OR 5.18, 95% CI: 2.98, 6.63; p<0.001) but not the PTV group (OR 1.17, 95% CI: 0.80-1.73; p=0.4) or the other missense variants or inframe deletion group (OR 0.99, 95% CI: 0.61, 1.61; p=1.0). Risk of ESKD per ICD was higher for Gly695Arg (OR 7.01, 95% CI: 3.48, 14.12; p<0.001), other collagenous domain glycine variants (OR 5.83, 95% CI: 1.67, 20.41; p=0.007), and PTVs (OR 3.43, 95% CI: 1.28, 9.19; p=0.02), compared to controls. Dipstick hematuria (trace blood or greater on at least 50% of urinalyses) was markedly increased for Gly695Arg (OR 9.47, 95% CI: 6.30, 14.22; p<0.001) and other glycine variants (OR 12.55, 95% CI: 6.02, 26.13), intermediate for PTVs (OR 1.63, 1.03, 2.58; 0.04), and not increased for other missense and inframe deletions (OR 0.62, 95% CI: 0.29, 1.33; p=0.2), compared to controls. The glycine collagenous domain variant groups also had worse KDIGO CKD risk categories than controls (**Figure 2, Table 2, Supplemental Figure 1**). Among patients with Gly695Arg, prevalence of high risk or greater KDIGO CKD risk category was 17.1% for those age 30-65 years and 53.3% for those 65+ years and older (**Figure 3, Supplemental Figure 2**).

**Figure 2.**
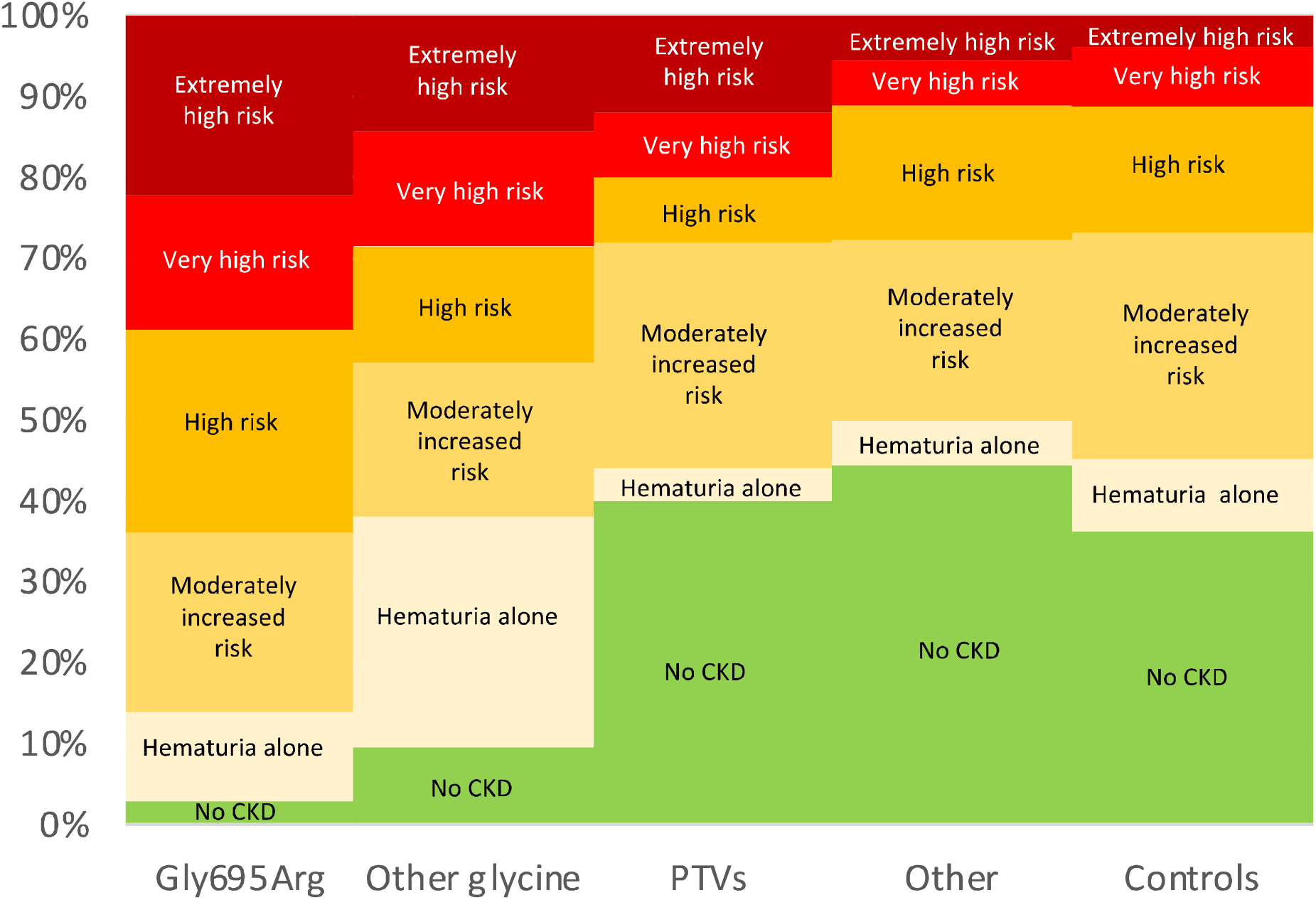
KDIGO CKD Risk Categories by Variant groups. KDIGO CKD Risk Categories: Hematuria only category had trace hematuria but eGFR>60 and ACR<30; moderately increased risk (eGFR>60 with ACR 30-300 or eGFR 45-59 with ACR<30); high risk (eGFR>60 with ACR >300, eGFR 45-59 with ACR 30-300, eGFR 30-44 with ACR<30); very high risk (eGFR 15-29 with ACR<30, eGFR 15-44 with ACR 30-300, eGFR 30-59 with ACR>300), extremely high risk (eGFR<15 or eGFR 15-29 with ACR>300, or ESKD by ICD code). If ACR data was unavailable, urinalysis data was used with dipstick 1+ twice classified as ACR 30-299 mg/g and dipstick 2+ twice or greater classified as ACR 300+ mg/g. This figure does not include any USRDS data.

**Figure 3.**
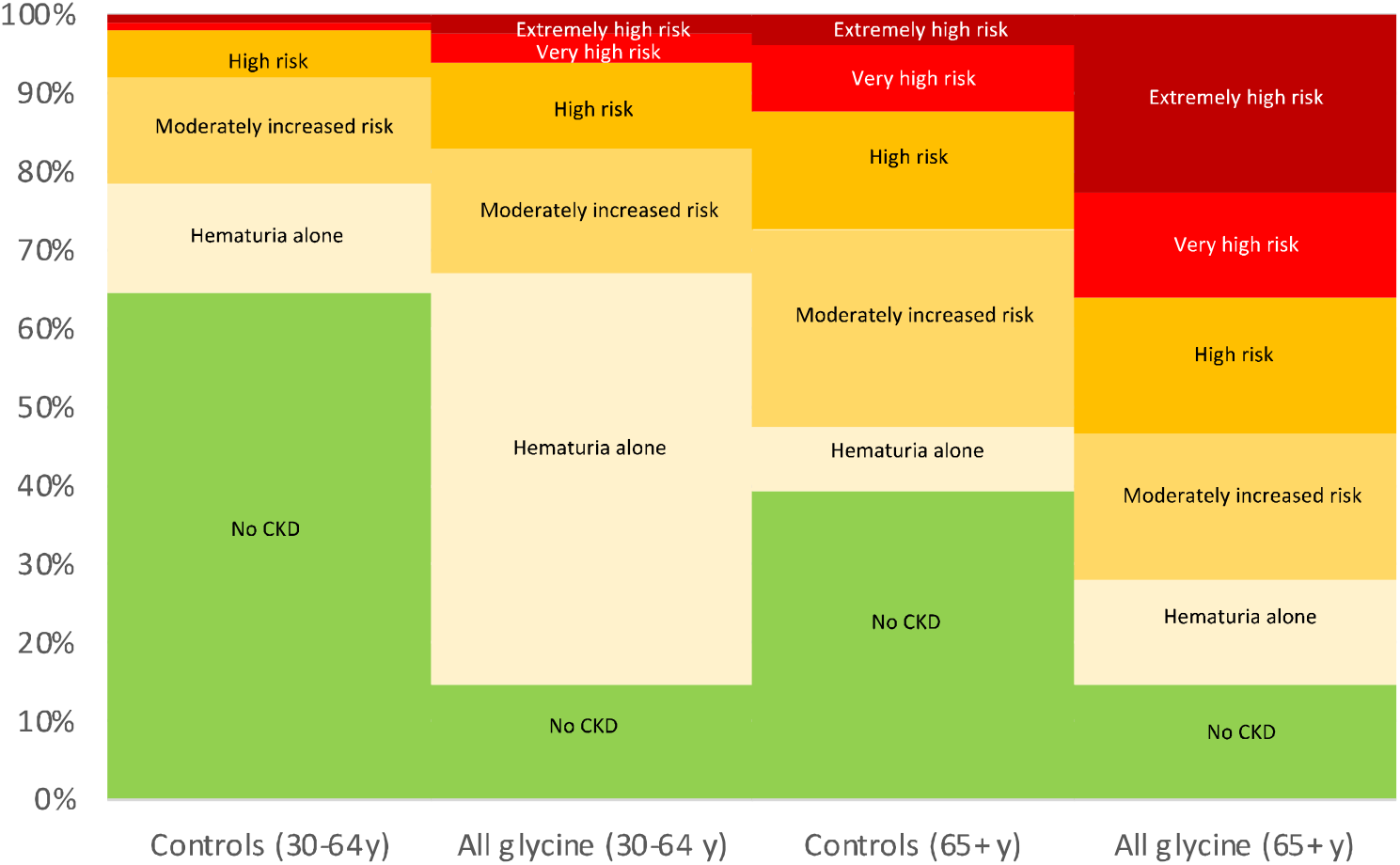
KDIGO CKD Risk Categories for Glycine Collagenous Domain Variants vs. Controls, by Age Group. This figure includes 169 patients heterozygous for glycine variants in the collagenous domain (including 129 Gly695Arg), and 1475 controls with eGFR and UA data. KDIGO CKD Risk Categories: Hematuria only category had trace hematuria but eGFR>60 and ACR<30; moderately increased risk (eGFR>60 with ACR 30-300 or eGFR 45-59 with ACR<30); high risk (eGFR>60 with ACR >300, eGFR 45-59 with ACR 30-300, eGFR 30-44 with ACR<30); very high risk (eGFR 15-29 with ACR<30, eGFR 15-44 with ACR 30-300, eGFR 30-59 with ACR>300), extremely high risk (eGFR<15 or eGFR 15-29 with ACR>300, or ESKD by ICD code). If ACR data was unavailable, urinalysis data was used with dipstick 1+ twice classified as ACR 30-299 mg/g and dipstick 2+ twice or greater classified as ACR 300+ mg/g. This figure does not include any USRDS data.

### Chart review of all COL4A3 P/LP heterozygotes

Only 4 patients had been diagnosed with AS or TBMD, and only 30% of patients were receiving ACE inhibitors or ARBs, the mainstay for treatment of AS (**Supplemental Table 5**). Nearly 1/3 of patients with glycine collagenous variants had undergone urologic workup for hematuria. Eleven patients underwent kidney biopsy; all had glycine collagenous domain variants and 4 (36.4%) had at least 1 other rare variant in a COL4A gene (**Table 4**). Diagnoses included: FSGS (n=6), TBMN (n=1), IgA nephropathy (n=2), light chain deposition disease (n=1), advanced glomerulosclerosis (n=1), and glomerulomegaly with severe tubular atrophy (n=1). Presence of thin GBM on kidney biopsy, a feature of ADAS was noted on 3 biopsies; one other biopsy noted variable GBM thickness ranging from 125-375 nm.

**Table 4.**
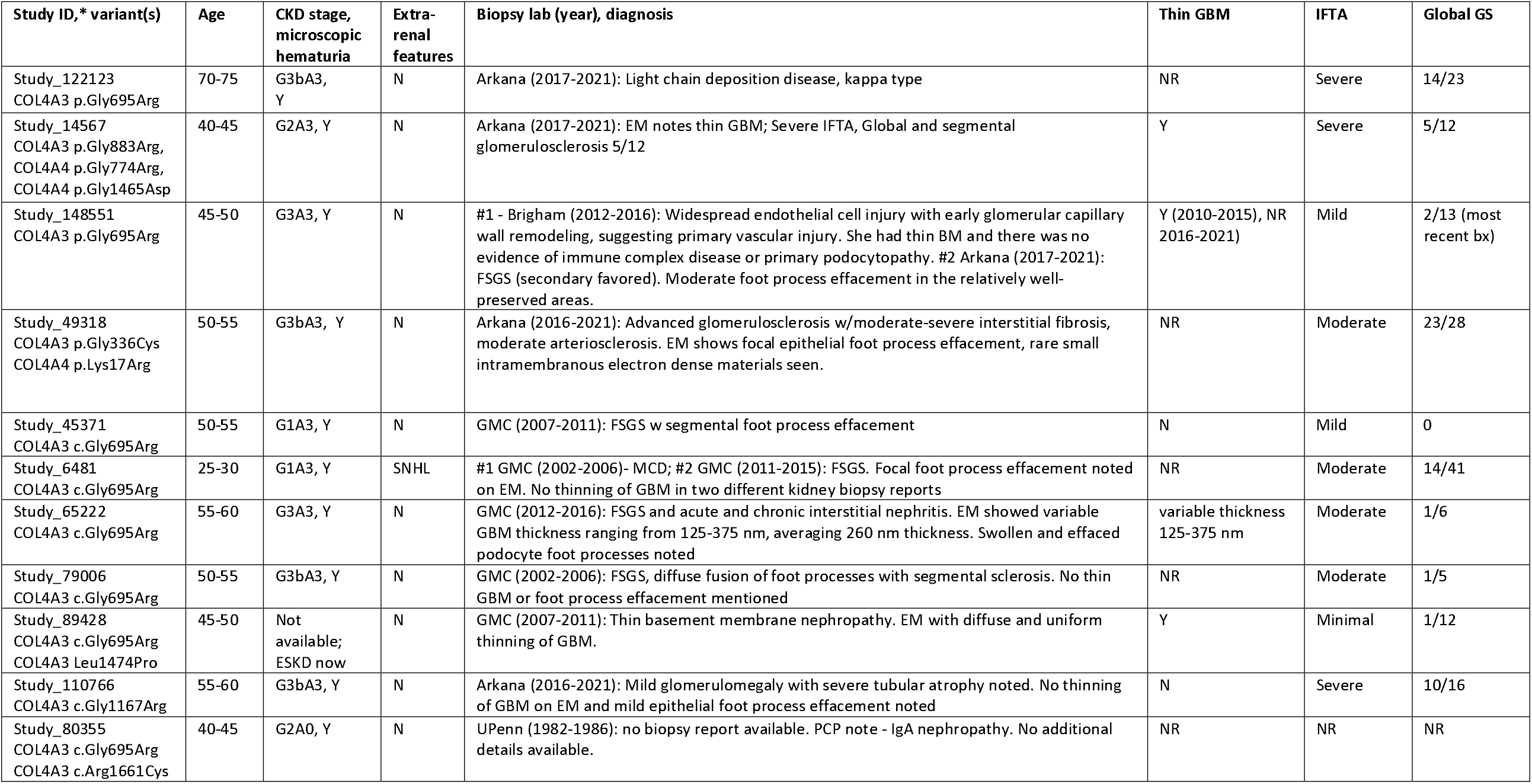
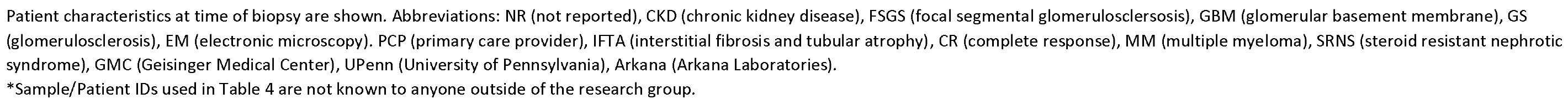
Characteristics of COL4A3 P/LP heterozygotes with kidney biopsies.

| Study ID,* variant(s) | Age | CKD stage, microscopic hematuria | Extra-renal features | Biopsy lab (year), diagnosis | Thin GBM | IFTA | Global GS |
| --- | --- | --- | --- | --- | --- | --- | --- |
| Study_122123<br>COL4A3 p.Gly695Arg | 70-75 | G3bA3, Y | N | Arkana (2017-2021): Light chain deposition disease, kappa type | NR | Severe | 14/23 |
| Study_14567<br>COL4A3 p.Gly883Arg,<br>COL4A4 p.Gly774Arg,<br>COL4A4 p.Gly1465Asp | 40-45 | G2A3, Y | N | Arkana (2017-2021): EM notes thin GBM; Severe IFTA, Global and segmental glomerulosclerosis 5/12 | Y | Severe | 5/12 |
| Study_148551<br>COL4A3 p.Gly695Arg | 45-50 | G3A3, Y | N | #1 - Brigham (2012-2016): Widespread endothelial cell injury with early glomerular capillary wall remodeling, suggesting primary vascular injury. She had thin BM and there was no evidence of immune complex disease or primary podocytopathy. #2 Arkana (2017-2021): FSGS (secondary favored). Moderate foot process effacement in the relatively well-preserved areas. | Y (2010-2015), NR (2016-2021) | Mild | 2/13 (most recent bx) |
| Study_49318<br>COL4A3 p.Gly336Cys<br>COL4A4 p.Lys17Arg | 50-55 | G3bA3, Y | N | Arkana (2016-2021): Advanced glomerulosclerosis w/moderate-severe interstitial fibrosis, moderate arteriosclerosis. EM shows focal epithelial foot process effacement, rare small intramembranous electron dense materials seen. | NR | Moderate | 23/28 |
| Study_45371<br>COL4A3 c.Gly695Arg | 50-55 | G1A3, Y | N | GMC (2007-2011): FSGS w segmental foot process effacement | N | Mild | 0 |
| Study_6481<br>COL4A3 c.Gly695Arg | 25-30 | G1A3, Y | SNHL | #1 GMC (2002-2006)- MCD; #2 GMC (2011-2015): FSGS. Focal foot process effacement noted on EM. No thinning of GBM in two different kidney biopsy reports | NR | Moderate | 14/41 |
| Study_65222<br>COL4A3 c.Gly695Arg | 55-60 | G3A3, Y | N | GMC (2012-2016): FSGS and acute and chronic interstitial nephritis. EM showed variable GBM thickness ranging from 125-375 nm, averaging 260 nm thickness. Swollen and effaced podocyte foot processes noted | variable thickness 125-375 nm | Moderate | 1/6 |
| Study_79006<br>COL4A3 c.Gly695Arg | 50-55 | G3bA3, Y | N | GMC (2002-2006): FSGS, diffuse fusion of foot processes with segmental sclerosis. No thin GBM or foot process effacement mentioned | NR | Moderate | 1/5 |
| Study_89428<br>COL4A3 c.Gly695Arg<br>COL4A3 Leu1474Pro | 45-50 | Not available;<br>ESKD now | N | GMC (2007-2011): Thin basement membrane nephropathy. EM with diffuse and uniform thinning of GBM. | Y | Minimal | 1/12 |
| Study_110766<br>COL4A3 c.Gly1167Arg | 55-60 | G3bA3, Y | N | Arkana (2016-2021): Mild glomerulomegaly with severe tubular atrophy noted. No thinning of GBM on EM and mild epithelial foot process effacement noted | N | Severe | 10/16 |
| Study_80355<br>COL4A3 c.Gly695Arg<br>COL4A3 c.Arg1661Cys | 40-45 | G2A0, Y | N | UPenn (1982-1986): no biopsy report available. PCP note - IgA nephropathy. No additional details available. | NR | NR | NR |

Patient characteristics at time of biopsy are shown. Abbreviations: NR (not reported), CKD (chronic kidney disease), FSGS (focal segmental glomerulosclerosis), GBM (glomerular basement membrane), GS (glomerulosclerosis), EM (electronic microscopy). PCP (primary care provider), IFTA (interstitial fibrosis and tubular atrophy), CR (complete response), MM (multiple myeloma), SRNS (steroid resistant nephrotic syndrome), GMC (Geisinger Medical Center), UPenn (University of Pennsylvania), Arkana (Arkana Laboratories).
\*Sample/Patient IDs used in Table 4 are not known to anyone outside of the research group.

Among the 19 patients with ESKD per USRDS, mean age of ESKD was 58.3 (13.4) years. Causes of ESKD listed on USRDS 2728 forms included AS (n=2), nephrotic syndrome with FSGS (n=3), as well as hypertension (n=2), renal artery stenosis (n=2), unspecified with renal failure (n=2), cholesterol emboli (n=2), type 2 diabetes, other vasculitis, IgA nephropathy, chronic interstitial nephritis, ADPKD, and unspecified injury.

## Discussion

In this study examining the phenotypic spectrum of pathogenic COL4A3 variants in an unselected population with a median of 15 years of follow-up time, we found that rare P/LP COL4A3 variant heterozygotes were at increased risks of hematuria, albuminuria, FSGS, CKD, and ESKD. Our results expand our knowledge of heterozygotes beyond previous studies mostly focusing on family members of patients. We observe that penetrance and phenotypic severity varied by genotype group with glycine missense variants located in the collagenous domain being particularly penetrant with at least 80% of individuals having any phenotypic feature. Among individuals with glycine variants in the collagenous domain and available data, roughly 1 out of 6 adults aged 30-65 years were KDIGO high risk or greater and more than 1 of 2 adults aged 65+ years were KDIGO high risk or greater. Over a median follow-up period of 15 years, the majority (70%) of the COL4A3 heterozygotes had never had ACR testing done before, highlighting the potential importance of early genetic diagnosis to inform appropriate management.

Prior studies have not been able to comprehensively examine penetrance of genetically-defined ADAS with varied reports depending on the study population (population-based vs. disease-specific) and the comprehensiveness of phenotyping. In case series of patients with ADAS or TBMN, the vast majority have hematuria, and most have proteinuria.^19–22^ In a systematic review of literature on TBMD due to COL4A3 or COL4A4,^21^ phenotypic spectrum of 777 patients with heterozygous COL4A3/COL4A4 mutations from 258 families included: 95% hematuria, 29% CKD, 15% ESKD (mean age ∼ 53 years), 16% hearing loss, 3% ocular lesions. However, the study noted lack of clear, uniform definitions for proteinuria, CKD, and hearing loss in the included studies, and did not compare these phenotypic features by COL4A3 variant groups. Results from research studies such as the Genomics England 100,000 Genomes Project report much lower prevalence of hematuria of 17.1% in collagenous domain glycine variants, due to reliance on defining hematuria from medical records.^15^ In our study with detailed phenotyping using extensive longitudinal EHR data including urinalyses, prevalence of hematuria was 70.3% using urinalysis data and 35.4% using ICD codes among patients heterozygous for COL4A3 Gly695Arg, a top hit for hematuria in genome-wide association studies. ^23 24^

We also confirmed a strong association between COL4A3 and FSGS,^25^ and found a prevalence of diagnosed FSGS of ∼2% in COL4A3 heterozygotes and ∼3% in Gly695Arg variant heterozygotes. This likely represents an underestimate as kidney biopsies are not done universally and many were not screened for albuminuria. In a study by Wang et al. examining rare variants in FSGS genes in 363 FSGS cases and 363 ancestry-matched controls, the top 3 contributors in a dominant or X-linked model were COL4A5, WT1, and COL4A4.^26^ The study also reported 8 COL4A3 variants in FSGS cases vs. 4 variants of unknown significance in matched controls; when restricted to glycine variants located in the collagenous domain, there were 6 in FSGS cases vs. 0 in controls. In a Toronto cohort of 193 individuals with FSGS from 174 families, 11% had a genetic diagnosis, including 28% in families with kidney disease, and 8% in sporadic cases. Heterozygous and hemizygous COL4A variants accounted for 55% of cases including 3 COL4A3 (all glycine variants located within the collagenous domain), 1 COL4A4, and 8 COL4A5.^5^

Among previously described COL4A3 P/LP variants, collagenous domain glycine missense variants had highest risk whereas PTVs had intermediate severity. Findings are consistent with previous literature that has shown that heterozygous COL4A3/COL4A4 pathogenic variants resulting in substitution of glycine residues in the collagenous domain with highly destabilizing residues (Arg, Val, Glu, Asp) were associated with increased risk of kidney failure and hematuria in comparison to those adjacent to non-collagenous domains.^27^ The weaker association of heterozygous PTVs compared to glycine collagenous domain variants has not been described previously to our knowledge and deserves further investigation. Type IV collagen proteins form protomers in the GBM, where α3(IV) forms multimers with α4(IV) and α5(IV).^28^ We hypothesize that disruption of the collagenous domain with glycine substitutions in one copy of COL4A3 will disrupt a significant proportion of heteromeric protein complexes, resulting in GBM defects and kidney phenotypes. In contrast, since the non-collagenous domains in the C-terminus of α3 chain of type IV collagen are essential to specificity and initiation of protomer assembly,^29,30^ PTVs of COL4A3 are likely impaired in heteromer formation. We hypothesize that for PTV carriers the reference (non-truncated) allele predominantly produces protein to form functional heteromers, which may be sufficient for basement membrane function, and therefore PTV carriers manifest a milder phenotype than carriers of glycine substitutions in the collagenous domain. In addition we found no association between other missense or inframe deletions that were not in the collagenous domain with kidney phenotypes although this group had higher prevalence of bilateral sensorineural hearing loss. Computational prediction programs to assign COL4A3/4/5 variant pathogenicity have been found to have limited value.^31^ Interestingly, out of 26 patients in our COL4A3 P/LP cohort who had an additional rare variant in a COL4A3/4/5 gene, 4 (15.4%) had ESKD, suggesting these second variants may contribute to AS severity. Further research will be needed to fully evaluate genotype-phenotype correlations, and individual variants’ pathogenicity.

Numerous opportunities for improved management of AS were identified, which may impact considerations for returning secondary findings of heterozygous COL4A3 P/LP variants to patients for earlier diagnosis and management.^32^ First, few patients with genetic ADAS had been diagnosed. Even among the patients with ESKD, only 2 were recognized as due to AS. Early knowledge could be important for ensuring ACR testing is done as patients at risk of CKD should undergo screening,^33^ and less than a third of COL4A3 heterozygotes had ACR testing. Three quarters of patients were not on ACEIs or ARBs, which have been shown to reduce risk of CKD progression in autosomal recessive AS with similar suggestive evidence in ADAS.^10,34 35^ In addition, knowledge of ADAS could be useful in minimizing unnecessary testing for hematuria as roughly a third of glycine collagenous domain variant heterozygotes underwent urologic workup of hematuria.

The major strengths of our study are the use of an unselected patient population with carefully phenotyping and use of propensity-matched controls over a median of 15 years follow-up, allowing us to provide insights on penetrance and phenotypic spectrum of ADAS. We were able to look at multiple phenotypic features of AS using a combination of ICD codes, longitudinal laboratory, USRDS, and kidney biopsy data. A limitation of the study is that we used EHR data rather than rigorously collected research data, which limits our ability to describe more subtle phenotypic features such as ocular manifestations. Our study population was mostly European in ancestry, and additional studies in other cohorts, including more diverse populations are needed. In conclusion we provide a strong foundation of evidence for the clinical significance and phenotypic spectrum of genetically-determined COL4A3 ADAS in an unselected population using carefully phenotyped EHR data while also demonstrating variability of phenotypic severity by genotype.

## Data Availability

The data supporting the findings of this study are available within the article and its Supplementary Data files. Additional information for reproducing the results described in the article is available upon reasonable request and subject to a data use agreement.

## Acknowledgements

We are grateful to the many patients who contributed to the MyCode Community Health Initiative by providing the genomic and electronic health information. We thank the Regeneron Genetics Center for providing funding for patient enrollment and exome sequencing for the DiscovEHR study, and we would like to acknowledge the Geisinger-Regeneron DiscovEHR Collaboration for the genotypic and phenotypic data. This work was supported by NIGMS grant GM111913 to T.M.

We thank the USRDS for their support. The data reported here have been supplied by the United States Renal Data System (USRDS). The interpretation and reporting of these data are the responsibility of the author(s) and in no way should be seen as an official policy or interpretation of the U.S. government.

## Author contributions

Study design (ARC, NTS), data collection (KS, BSM, TM, VA, VA), analysis (YH, ARC), interpretation of results (KS, NTS, IDB, TM, NTS, ARC), drafted manuscript (KS, ARC), revised critically for important intellectual content (all authors). All authors have approved the version to be published.

## Ethics Declaration

This research was approved by the Geisinger Clinic Institutional Review Board and included participants in the MyCode Health Initiative who have exome sequencing data obtained as part of the Geisinger-Regeneron DiscovEHR collaboration. All participants provided written informed consent, and all experiments were performed in accordance with relevant guidelines and regulations.

