## Supplemental Materials for "The Phenotypic Spectrum of *COL4A3* Heterozygotes"

**Supplemental Methods.**

**Supplemental Table 1. Coding definitions for comorbidities**

**Supplemental Table 2. Characteristics by individual variants**

**Supplemental Table 3. Characteristics of COL4A3 P/LP Heterozygotes and non-heterozygotes before and after matching**

**Supplemental Table 4. *COL4A3* P/LP heterozygotes with an additional rare variant in *COL4A3/4/5***

**Supplemental Table 5. Chart Review data of *COL4A3* AD Alport Syndrome-related Diagnosis and Management**

**Supplemental Methods.**

Sequencing was performed by paired end 75bp reads on either an Illumina HiSeq2500 or NovaSeq. Coverage depth was sufficient to provide more than 20% coverage over 85% of the targeted bases in 96% of the VCR samples and 90% coverage for 99% of IDT samples. Alignments and variant calling were based on GRCh38 human genome reference sequence. We required AD_ALT>3 and balance AD_ALT/AD_REF ≥0.5.

**Supplemental Table 1. Coding definitions for comorbidities**

| **Comorbidities** | **Codes** |
| --- | --- |
| Hematuria | ICD-10: R31.0, R31.1, R31.21, R31.29, R31.9; ICD-9: 599.7* |
| FSGS | ICD-10: N03.1, N04.1, N05.1, N06.1; ICD-9: 581.1 |
| Glomerulonephritis | ICD-10: N00.9, N0.39, N05.8, N02.2, N05.5, N05.9, N02.8, N05.9; ICD-9: 580.9, 582.9 |
| Hearing loss | H90.* |
| Sensorineural hearing loss | ICD-10: H90.3; ICD-9: 389.11, 389.12, 389.14, 389.18 |
| ESKD | Dialysis ICD codes 39.27, 39.42, 39.53, 39.54, 585.6, V45.11, V45.12, V56.1, V56.2, V56.31, V56.32, V56.8, V45.1, N18.6, Z91.15, N18.5+Z99.2 Transplant ICD codes 00.91, 00.92, 00.93, 55.53, 55.69, V42.0, 0TY****, Z94.0 |
| Hypertension | ICD-9: 401 - 405 |
|  | ICD-10: I10 - I16 |
|  | ICD-10: E78 |
| Diabetes mellitus | ICD-9: 250 |
|  | ICD-10: E10, E11, E13 |

**Supplemental Table 2. Characteristics by individual variants**

| **Variant** | **ClinVar (as of 2/23/23)** | **Varsome ACMG Classifier (as of 2/23/23)** | **Age** | **Hematuria ICD code** | **Hearing loss ICD code** | **eGFR <60** | **eGFR <30** | **ESKD ICD code** | **UA available** | **Trace blood on 50%** | **1+ blood on 50%** |
| --- | --- | --- | --- | --- | --- | --- | --- | --- | --- | --- | --- |
| **rs1014839148 (n = 2)** 2: 227248494:G:A c.520G>A p.Gly174Arg | Conflicting (1 pathogenic, 1 likely pathogenic, 2 uncertain significance); 4 submissions; 1 star | Likely pathogenic (9 points) | 80.7 (SD 13.8) | 50% | 50% | 50% | 0 | 0 | 100% | 100% | 100% |
| **rs1057516204 (n=1)** 2:227293218:GGAAA:G c.3244_3247del p.Lys1082fs | Pathogenic; 2 submissions; 1 star | Pathogenic (10 points) | 74.2 | 0% | 0 | 100% | 0 | 0 | 100% | 0 | 0 |
| **rs1064796094 (n=2)** 2:227290085:CCA:C c.3068_3069del p.Pro1023fs | Pathogenic/Likely pathogenic; 3 submissions; 2 stars | Pathogenic (17 points) | 54 (SD 18.4) | 50% | 0 | 0 | 0 | 0 | 100% | 100% | 50% |
| **rs1158937060 (n=5)** 2:227273119:T:C c.1927+2T>C | Likely pathogenic; 2 submissions; 2 stars | Pathogenic (11 points) | 55.6 (SD 21.2) | 0 | 0 | 33.30% | 0 | 0 | 100% | 0 | 0 |
| **rs1167411352 ( n=1)** 2:227297727:G:C c.3619G>C Gly1207Arg | Pathogenic; 1 ssubmission; 1 star | Pathogenic (16 points) | 48.8 | 0 | 0 | 0 | 0 | 0 | 100% | 100% | 0 |
| **rs1173685095 (n=9)** 2:227290090:T:C c.3070+2T>C | Likely pathogenic; 1 submission; 0 stars | Pathogenic (10 points) | 60.0 (SD 21.2) | 33.30% | 0 | 37.50% | 25% | 22.20% | 77.80% | 85.70% | 71.40% |
| **rs1175052474 (n=12)** 2:227295295:A:AGAG c.3546_3548dup p.Gly1183dup | Conflicting (2 pathogenic, 2 likely pathogenic, 2 uncertain significance); 9 submissions; 1 star | Likely pathogenic (6 points) | 65.8 (SD 12.4) | 50% | 0.00% | 50% | 8.30% | 0.00% | 83.30% | 90% | 70% |
| **rs1192750535 (n=1)** 2:227293230:G:T c.3250G>T p.Glu1084Ter | Pathogenic; 2 submissions; 2 stars | Pathogenic (13 points) | 39.5 | 0 | 0 | 0 | 0 | 0 | 100% | 100% | 0 |
| **rs1196996393 (n=1)** 2:227284210:G:C c.2747-1G>C | Pathogenic; 2 submissions; 2 stars | Pathogenic (13 points) | 73.6 | 0.0% | 0.0% | 0.0% | 0.0% | 0.0% | 0.0% | N/A | N/A |
| **rs756539994 (n=21)** 2:227293286:CCCTGGAAGT:C c.3312_3329del | Conflicting (2 pathogenic, 1 likely pathogenic, 3 uncertain significance); 4 submissions; 1 star | Likely pathogenic (6 points) | 56.3 (SD 15.7) | 14.3% | 14.3% | 10.5% | 5.3% | 0.0% | 76.2% | 18.8% | 18.8% |
| **rs759043857 (n=20)** 2:227282495:AG:A c.2621del p.Gly874fs | Likely pathogenic; 3 submissions; 2 stars | Pathogenic (17 points) | 57.6 (SD 20.6) | 15.0% | 10.0% | 23.5% | 0.0% | 0.0% | 65.0% | 7.7% | 7.7% |
| **rs760462252 (n=39**) 2:227307839:C:T c.4382C>T p.Pro1461Leu | Pathogenic; 1 submission; 1 star | Uncertain significance (5 points) | 56.6 (SD 20.5) | 10.3% | 15.4% | 19.4% | 2.8% | 2.6% | 84.6% | 12.1% | 9.1% |
| **rs760846085 (n=32)** 2:227310822:CT:C c.4803del p.Gly1602fs | Pathogenic/likely pathogenic; 4 submissions; 2 stars | Pathogenic (17 points) | 61.6 (SD 17.7) | 12.5% | 0.0% | 25.0% | 7.1% | 6.3% | 87.5% | 28.6% | 17.9% |
| **rs868002181 (n=10)** 2:227280970:G:A c.2452G>A p.Gly818Arg | Conflicting (2 pathogenic, 1 likely pathogenic, 1 uncertain significance); 6 submissions; 1 star | Likely pathogenic (7 points) | 63.0 (SD 14.0) | 30.0% | 0.0% | 30.0% | 20.0% | 0.0% | 90.0% | 62.5% | 50.0% |
| **rs200287952 (n=161)** 2:227277511:G:A 2083G>A Gly695Arg | Pathogenic; 3 submissions; 1 star | Likely pathogenic (8 points) | 59.4 (SD 19.1) | 35.4% | 4.4% | 31.7% | 12.4% | 8.1% | 79.5% | 70.3% | 55.5% |
| **rs121912824 (n=3)** 2:227307897:C:T c.4441C>T p.Arg1481Ter | Pathogenic; 7 submissions; 2 stars | Pathogenic (17 points) | 56.5 (SD 28.4) | 0.0% | 0.0% | 0.0% | 0.0% | 0.0% | 66.6% | 0.0% | 0.0% |
| **rs1306992119 (n=1)** 2:227280529:ACTCCCTGGACTTCCAGGT:A c.2323_2340del | Pathogenic/Likely pathogenic; 3 submissions; 2 stars | Likely pathogenic (7 points) | 51.5 | 0.0% | 0.0% | 0.0% | 0.0% | 0.0% | 66.7% | 0.0% | 0.0% |
| **rs1445615417 (n=2)** 2:227307867:GTTTTC:G c.4420_4424del p.Leu1474fs | Pathogenic; 4 submissions; 2 stars | Pathogenic (17 points) | 52.0 (SD 22.3) | 50.0% | 0.0% | 0.0% | 0.0% | 0.0% | 100.0% | 0.0% | 0.0% |
| **rs1453590085 (n=1)** 2:227164754:C:T c.28C>T p.Gln10Ter | Pathogenic; 2 submissions; 2 stars | Pathogenic (13 points) | 39 | 0.0% | 0.0% | 0.0% | 0.0% | 0.0% | 0.0% | N/A | N/A |
| **rs1469479748 (n=1)** 2:227253310:AAG:A c.663_664del  p.Arg221fs | Pathogenic/likely pathogenic; 4 submissions; 2 stars | Pathogenic (17 points) | 64.9 | 0.0% | 0.0% | 0.0% | 0.0% | 0.0% | 100.0% | 0.0% | 0.0% |
| **rs1553751120 (n=1)** 2:227247583:AG:A c.468+1del | Likely pathogenic; 1 submission; 1 star | Pathogenic (11 points) | 62.2 | 0.0% | 0.0% | 0.0% | 0.0% | 0.0% | 0.0% | N/A | N/A |
| **rs1553751122(n=5)** 2:227247585:G:T c.468+1G>T | Likely pathogenic; 1 submission; 1 star | Pathogenic (11 points) | 60.8 (SD 20.0) | 20.0% | 0.0% | 60.0% | 20.0% | 0.0% | 80.0% | 75.0% | 25.0% |
| **rs1553766404 (n=3)** 2: 227309077:G:T c.4640+1G>A | Likely pathogenic; 1 submission; 1 star | Pathogenic (11 points) | 53.5 (SD 17.5) | 33.3% | 0.0% | 33.3% | 0.0% | 0.0% | 100.0% | 0.0% | 0.0% |
| **rs1559854632 (n=2)** 2: 227237968 c.88G>C Gly30Arg | Likely pathogenic; 1 submission; 0 stars | Uncertain significance (2 points) | 62.3 (SD 16.3) | 50.0% | 0.0% | 50.0% | 0.0% | 0.0% | 100.0% | 50.0% | 0.0% |
| **rs 1559873550 (n=1)** 2: 227257621:G:T c.1006G>T p.Gly336Cys | Pathogenic/likely pathogenic; 4 submissions; 2 stars | Pathogenic (14 points) | 55.3 | 0.0% | 0.0% | 100.0% | 0.0% | 100.0% | 100.0% | 100.0% | 100.0% |
| **rs1559878824 (n=2)** 2: 227263830:G:A c.1201G>A p.Gly401Arg | Conflicting (1 likely pathogenic, 1 uncertain significance); 3 submissions; 1 star | Likely pathogenic (6 points) | 55.7 (SD 9.0) | 50.0% | 50.0% | 50.0% | 0.0% | 0.0% | 100.0% | 50.0% | 50.0% |
| **rs1559897288 (n=2)** 2: 227282523:G:A c.2647G>A p.Gly883Arg | Likely pathogenic; 1 submission; 0 stars | Likely pathogenic (7 points) | 45.4 (SD 1.5) | 100.0% | 50.0% | 0.0% | 0.0% | 0.0% | 100.0% | 100.0% | 100.0% |
| **rs1574658390 (n=2)** 2: 227240203:G:T c.205G>T p.Glu69Ter | Likely pathogenic; 1 submission; 0 stars | Pathogenic (10 points) | 78.9 (SD 15.5) | 100.0% | 0.0% | 100.0% | 0.0% | 0.0% | 50.0% | 100.0% | 0.0% |
| **rs200672668 (n=2)** 2: 227273108:G:A c.1918G>A p.Gly640Arg | Pathogenic/likely pathogenic; 6 submissions; 2 stars | Pathogenic (18 points) | 49.0 (SD 19.4) | 50.0% | 0.0% | 0.0% | 0.0% | 0.0% | 50.0% | 100.0% | 0.0% |
| **rs202147112 (n=4**) 2: 227245972:G:A c.343G>A p.Gly115Arg | Conflicting (2 likely pathogenic, 1 uncertain significance); 4 submissions; 1 star | Likely pathogenic (8 points) | 44.8 (SD 12.02) | 0.0% | 0.0% | 0.0% | 0.0% | 0.0% | 100.0% | 25.0% | 25.0% |
| **rs267606745 (n=3)** 2: 227295044:G:A c.3499G>A p.Gly1167Arg | Pathogenic; 6 submissions; 2 stars | Pathogenic (20 points) | 44.1 (SD 15.1) | 100.0% | 0.0% | 50.0% | 0.0% | 0.0% | 100.0% | 100.0% | 100.0% |
| **rs368434069 (n=4)** 2: 227297751:C:T c.3643C>T p.Arg1215Ter | Pathogenic; 3 submissions; 2 stars | Pathogenic (17 points) | 41.3 (SD 18.3) | 50.0% | 0.0% | 0.0% | 0.0% | 0.0% | 100.0% | 0.0% | 0.0% |
| **rs371334239 (n=5)** 2:227263845:C:T c.1216C>T p.Arg406Ter | Pathogenic; 6 submissions; 2 stars | Likely benign (-4 points) | 48.9 (SD 23.1) | 20.0% | 0.0% | 0.0% | 0.0% | 0.0% | 100.0% | 60.0% | 20.0% |
| **rs375040636 (n=1)** 2: 227279882:G:A c.2215G>A p.Gly739Arg | Likely pathogenic; 3 submissions; 2 stars | Pathogenic (20 points) | 72.5 | 0.0% | 0.0% | 100.0% | 0.0% | 0.0% | 100.0% | 100.0% | 100.0% |
| **rs573527081 (n=6)** 2: 227253637:C:T 764C>T Thr255Met | Conflicting (1 likely pathogenic; 2 uncertain significance); 4 submissions; 1 star | Uncertain significance (2 points) | 63.4 (SD 28.2) | 0.0% | 16.7% | 0.0% | 0.0% | 0.0% | 66.7% | 0.0% | 0.0% |
| **rs748026887 (n=2)** 2:227307800:TCACCCGA:T c.4347_4353del p.Arg1450fs | Pathogenic; 5 submissions; 2 stars | Pathogenic (17 points) | 73.3 (SD 15.9) | 0.0% | 0.0% | 0.0% | 0.0% | 0.0% | 50.0% | 0.0% | 0.0% |
| **rs766208466 (n=6)** 2: 227310803:G:A c.4783G>A p.Gly1595Arg | Conflicting (1 likely pathogenic, 1 uncertain significance); 3 submissions; 1 star | Uncertain significance (5 points) | 57.6 (SD 15.9) | 0.0% | 0.0% | 20.0% | 0.0% | 0.0% | 83.3% | 0.0% | 0.0% |
| **rs766306957 (n=8)** 2:227284229:GAGTAAAGGGCC:G c.2768_2778del | Pathogenic; 6 submissions; 2 stars | Pathogenic (17 points) | 65.5 (SD 16.4) | 37.5% | 0.0% | 0.0% | 0.0% | 0.0% | 100.0% | 12.5% | 12.5% |
| **rs766900945 (n=2)** 2:227290785:C:T  c.3109C>T p.Arg1037Ter | Pathogenic; 5 stars; 2 stars | Likely benign (-2 points) | 81.2 (SD 13.2) | 50.0% | 0.0% | 50.0% | 50.0% | 0.0% | 100.0% | 50.0% | 0.0% |
| **rs769683665 (n=3)** 2:227289230:G:A c.2962G>A p.Gly988Arg | Likely pathogenic; 1 submission; 0 stars | Pathogenic (10 points) | 79.4 (SD 9.8) | 33.3% | 0.0% | 33.3% | 0.0% | 0.0% | 66.7% | 0.0% | 0.0% |
| **rs769863513 (n=4)** 2:227308922:C:T c.4486C>T p.Arg1496Ter | Pathogenic/likely pathogenic; 4 submissions; 2 stars | Pathogenic (17 points) | 61.3 (SD 14.8) | 0.0% | 25.0% | 0.0% | 0.0% | 0.0% | 50.0% | 0.0% | 0.0% |
| **rs779575469 (n=1**) 2:227270788:G:T c.1594G>T p.Gly532Cys | Pathogenic; 2 submissions; 1 star | Pathogenic (14 points) | 51.8 | 0.0% | 0.0% | 0.0% | 0.0% | 0.0% | 100.0% | 100.0% | 100.0% |
| **rs867868993 (n=1)** 2:227310818:CCCC:CCC c.4802del p.Pro1601fs | Pathogenic/Likely pathogenic; 2 submissions; 2 stars | Pathogenic (11 points) | 75.4 | 0.0% | 0.0% | 0.0% | 0.0% | 0.0% | 100.0% | 0.0% | 0.0% |
| **rs914878176 (n=3)** 2:227295016:G:C c.3472G>C p.Gly1158Arg | Conflicting (1 pathogenic, 2 likely pathogenic, 1 uncertain significance); 5 submissions; 1 star | Likely pathogenic (7 points) | 43.3 (SD 5.5) | 33.3% | 0.0% | 0.0% | 0.0% | 0.0% | 100.0% | 100.0% | 66.7% |
| **rs988439345 (n=2)** 2:227253581:AC:A c.713del p.Pro240fs | Pathogenic; 1 submission; 1 star | Pathogenic (11 points) | 69.5 (SD 8.3) | 0.0% | 0.0% | 100.0% | 0.0% | 0.0% | 100.0% | 50.0% | 50.0% |
| **rs993103826 (n=2)** 2:227282410:GC:G c.2535del p.Leu846fs | Pathogenic/Likely pathogenic; 3 submissions; 2 stars | Pathogenic (17 points) | 39.1 (SD 6.3) | 0.0% | 0.0% | 0.0% | 0.0% | 0.0% | 100.0% | 50.0% | 50.0% |

**Supplemental Table 3. Characteristics of *COL4A3* P/LP Heterozygotes and non-heterozygotes before and after matching**

|  | **Before matching (N=174361)*** | |  |  | **After propensity score matching (N=2411)** | |  |  |
| --- | --- | --- | --- | --- | --- | --- | --- | --- |
|  | **COL4A3 P/LP heterozygotes (n=402)** | **Non-heterozygotes (n=173959)** | **P value** | **SMD** | **COL4A3 P/LP heterozygotes (n=402)** | **Non-heterozygotes (n=2009)** | **P value** | **SMD** |
| Age, Mean (SD) | 59.0 (18.6) | 57.5 (18.9) | 0.104 | 0.082 | 59.1 (18.7) | 59.1 (18.5) | 0.986 | 0.001 |
| Female (%) | 257 (63.9) | 105441 (60.6) | 0.19 | 0.069 | 257 (63.9) | 1284 (63.9) | 1 | <0.001 |
| Black, n (%) | 9 (2.2) | 3951 (2.3) | 1 | 0.002 | 9 (2.2) | 32 (1.6) | 0.482 | 0.047 |
| Hispanic, n (%) | 4 (1.0) | 4613 (2.7) | 0.056 | 0.124 | 4 (1.0) | 47 (2.3) | 0.128 | 0.105 |
| Year of first outpatient visit, median (IQI) | 2003 (2001-2011) | 2004 (2001-2011) | 0.12 | 0.079 | 2003 (2001-2011) | 2003 (2001-2011) | 0.939 | 0.004 |
| Hypertension (%) | 226 (56.2) | 92964 (53.3) | 0.259 | 0.059 | 226 (56.2) | 1129 (56.2) | 1 | <0.001 |
| Diabetes (%) | 90 (22.4) | 41056 (23.6) | 0.608 | 0.029 | 90 (22.4) | 449 (22.3) | 1 | 0.001 |
| Nephrolithiasis (%) | 22 (5.5) | 10155 (5.8) | 0.838 | 0.016 | 22 (5.5) | 109 (5.4) | 1 | 0.002 |
| Hematuria, n (%) | 110 (27.4) | 23404 (13.5) | <0.001 | 0.35 | 110 (27.4) | 279 (13.9) | <0.001 | 0.338 |
| FSGS, n (%) | 7 (1.7) | 437 (0.3) | <0.001 | 0.15 | 7 (1.7) | 4 (0.2) | <0.001 | 0.158 |
| Bilateral sensorineural hearing loss, n (%) | 23 (5.7) | 8340 (4.8) | 0.453 | 0.042 | 23 (5.7) | 109 (5.4) | 0.906 | 0.013 |
| eGFR available, n (%) | 358 (89.1) | 154420 (88.8) | 0.918 | 0.009 | 358 (89.1) | 1815 (90.3) | 0.484 | 0.042 |
| eGFR mean (SD) | 74.9 (28.8) | 80.9 (26.6) | <0.001 | 0.214 | 74.9 (28.8) | 80.3 (27.1) | 0.001 | 0.192 |
| eGFR <60, n (%) | 97/360 (26.9) | 31512/154586 (20.4) | 0.003 | 0.155 | 97/360 (26.9) | 388/1817 (21.4) | 0.024 | 0.131 |
| eGFR <30, n (%) | 30/360 (8.3) | 5551/154586 (3.6) | <0.001 | 0.201 | 30/360 (8.3) | 59/1817 (3.3) | <0.001 | 0.219 |
| Urinalysis available, n(%) | 326 (81.1) | 131667 (75.7) | 0.014 | 0.132 | 326 (81.1) | 1561 (77.7) | 0.15 | 0.084 |
| Trace or greater, n (%) | 158/326 (48.5) | 28166/131667 (21.4) | <0.001 | 0.592 | 158/326 (48.5) | 327/1561 (21.0) | <0.001 | 0.604 |
| 1+ or greater, n (%) | 119/326 (36.5) | 16508/131667 (12.5) | <0.001 | 0.58 | 119/326 (36.5) | 184/1561 (11.8) | <0.001 | 0.603 |
| ESKD per USRDS, n (%) | 19 (4.7) | 1827 (1.1) | <0.001 | 0.187 | 19 (4.7) | 18 (0.9) | <0.001 | 0.194 |
| ESKD per ICD code, n (%) | 23 (5.7) | 2668 (1.5) | <0.001 | 0.225 | 23 (5.7) | 25 (1.2) | <0.001 | 0.246 |
| ESKD per USRDS or ICD code, n (%) | 23 (5.7) | 2862 (1.7) | <0.001 | 0.218 | 23 (5.7) | 27 (1.3) | <0.001 | 0.239 |
| ACR available, n (%) | 118 (29.4) | 48836 (28.1) | 0.607 | 0.028 | 118 (29.4) | 539 (26.8) | 0.329 | 0.056 |
| ACR >30, n (%) | 50/118 (42.4) | 11990/48836 (24.6) | <0.001 | 0.385 | 50/118 (42.4) | 133/539 (24.7) | <0.001 | 0.382 |
| ACR >300, n (%) | 20/118 (17.0) | 2692/48836 (5.5) | <0.001 | 0.368 | 20/118 (17.0) | 29/538 (5.4) | <0.001 | 0.374 |
| At least 1 phenotypic feature^ | 242 (60.2) | 76311 (38.7) | <0.001 | 0.44 | 242 (60.2) | 797 (39.7) | 1 | 0.419 |

*Whole Exome Sequencing (WES): WES was performed by paired end 75bp reads on either an Illumina HiSeq2500 or NovaSeq. Coverage depth was sufficient to provide more than 20% coverage over 85% of the targeted bases in 96% of the VCR samples and 90% coverage for 99% of IDT samples. Alignments and variant calling were based on GRCh38 human genome reference sequence. We required AD_ALT>3 and balance od AD_ALT/AD_REF ≥0.5.

| **Study ID** | **COL4A3 P/LP variant** | **Additional heterozygous rare variant(s) in COL4A3/4/5; ACMG classification per Varsome** | **Phenotype** |
| --- | --- | --- | --- |
| study_108043 | p.Gly30Arg | COL4A4 p.Thr1474Met (LB; -1 points) | Stage 3a CKD. No dipstick hematuria, FSGS, bilateral sensorineural hearing loss, ESKD. Missing ACR data. |
| study_111302 | c.468+1del | COL4A4 p.Pro1587Arg (B; -8 points) | No CKD. No FSGS, bilateral sensorineural hearing loss, ESKD. Missing UA and ACR data. |
| Study_130305 | p.Gly818Arg | COL4A4 p.Pro1587Arg (B; -8 points) | ESKD, eGFR<15, +dipstick hematuria, + severe albuminuria. No FSGS or bilateral sensorineural hearing loss. |
| Study_130349 | p.Gly695Arg | COL4A4 p.Pro1587Arg (B; -8 points) | No CKD. No FSGS, bilateral sensorineural hearing loss, ESKD. Missing UA and ACR data. |
| Study_143022 | p.Gly818Arg | COL4A4 p.Pro1587Arg (B; -8 points) | Hematuria alone. No FSGS, bilateral sensorineural hearing loss, albuminuria, or ESKD. |
| Study_14567 | p.Gly695Arg | COL4A4 p.Gly774Arg (VUS; 5 points) COL4A4 p.Gly1465Asp (VUS; 3 points) | Hematuria + severe albuminuria, eGFR >60. |
| Study_146959 | p.Gly115Arg | COL4A3 p.Arg341His (VUS; 1 point) | No CKD. No dipstick hematuria, FSGS, bilateral sensorineural hearing loss, or ESKD. Missing ACR data. |
| Study_149658 | p.Gly874AspfsTer9 | COL4A4 p.Arg877Gln (B; -12 points) | No CKD. No dipstick hematuria, FSGS, bilateral sensorineural hearing loss, or ESKD. Missing ACR data. |
| Study_151405 | p.Gly818Arg | COL4A4 p.Pro1587Arg (B; -8 points) | No CKD. No dipstick hematuria, FSGS, bilateral sensorineural hearing loss, or ESKD. Missing ACR data. |
| Study_152777 | c.468+1G>T | COL4A3 p.Val1560Ile (LB; -3 points) | Stage 3a CKD with severe albuminuria and dipstick hematuria. No FSGS, bilateral sensorineural hearing loss, or ESKD. |
| Study_15491 | p.Gly818Arg | COL4A4 p.Pro1587Arg (B; -8 points) | No CKD. No dipstick hematuria, FSGS, bilateral sensorineural hearing loss, or ESKD. Missing ACR data |
| Study_158376 | p.Gly640Arg | COL4A4 p.Gln1473Lys (VUS; 4 points) | No CKD. No FSGS, bilateral sensorineural hearing loss, or ESKD. Missing ACR and UA data. |
| Study_1600 | p.Gly818Arg | COL4A4 p.Pro1587Arg (B; -8 points) | Stage 3a CKD. No FSGS, bilateral sensorineural hearing loss, or ESKD. Missing ACR and UA data. |
| Study_42724 | p.Gly695Arg | COL4A4 p.Arg877Gln (B; -12 points) | Hematuria alone. eGFR>60, no albuminuria, FSGS, bilateral sensorineural hearing loss, or ESKD. |
| Study_46348 | c.3070+2T>C | COL4A3 p.Leu1474Pro (VUS; 3 points) | ESKD + hematuria. No FSGS, bilateral sensorineural hearing loss. Missing ACR data |
| Study_48927 | p.Gly818Arg | COL4A4 p.Pro1587Arg (B; -8 points) | Hematuria alone. eGFR>60, no albuminuria, FSGS, bilateral sensorineural hearing loss, or ESKD. |
| Study_49318 | p.Gly336Cys | COL4A4 p.Lys17Arg (B; -13 points) | ESKD + Hematuria + mild albuminuria. No FSGS, bilateral sensorineural hearing loss. |
| Study_5112 | p.Gly818Arg | COL4A4 p.Pro1587Arg (B; -8 points) | Hematuria only. eGFR>60, no FSGS, bilateral sensorineural hearing loss, or ESKD. Missing ACR data |
| Study_55019 | p.Gly695Arg | COL4A4 p.Pro1587Arg (B; -8 points) | Hematuria alone. eGFR>60, no FSGS, bilateral sensorineural hearing loss, or ESKD. Missing ACR data |
| Study_61689 | p.Gly818Arg | COL4A4 p.Pro1587Arg (B; -8 points) | No CKD. No FSGS, bilateral sensorineural hearing loss, or ESKD. Missing ACR and UA data. |
| Study_66784 | p.Gly1602AlafsTer13 | COL4A3 p.Asn1508Ser (VUS; 0 points) | Hematuria alone. eGFR>60, no FSGS, bilateral sensorineural hearing loss, or ESKD. Missing ACR data |
| Study_69681 | p.Gly818Arg | COL4A4 p.Pro1587Arg (B; -8 points) | Hematuria alone. eGFR>60, no FSGS, bilateral sensorineural hearing loss, or ESKD. Missing ACR data |
| Study_77870 | p.Gly30Arg | COL4A4 p.Thr1474Met (LB; -1 points) | Hematuria alone. eGFR>60, no albuminuria, FSGS, bilateral sensorineural hearing loss, or ESKD. |
| Study_80355 | p.Gly695Arg | COL4A3 p.Arg1661Cys (LP; 9 points) | Missing eGFR, ACR, and UA data. No FSGS, bilateral sensorineural hearing loss, or ESKD. |
| Study_87695 | p.Gly874AspfsTer9 | COL4A3 p.Leu1474Pro (VUS; 3 points) | No CKD. No FSGS, bilateral sensorineural hearing loss, or ESKD. Missing ACR and UA data. |
| Study_89428 | p.Gly695Arg | COL4A3 p.Leu1474Pro (VUS; 3 points) | ESKD + hematuria + bilateral sensorineural hearing loss. No FSGS. Missing ACR data. |

**Supplemental Table 4. *COL4A3* P/LP heterozygotes with an additional rare variant in *COL4A3/4/5***

This table does not use USRDS data and Sample/Patient IDs are not known to anyone outside of the research group

**Supplemental Table 5. Chart Review data of *COL4A3* AD Alport Syndrome-related Diagnosis and Management**

| **Variables n, (%)** | **Prevalence of P/LP overall *COL4A3* variants and their subgroups** | | | | |
| --- | --- | --- | --- | --- | --- |
|  | **Overall (n=402)** | **Gly695Arg (n=161)** | **Other collagenous domain glycine variants**  **(n=47)** | **PTVs (n=119)** | **Other missense or inframe deletions**  **(n=75)** |
| **Diagnosed with Alport Syndrome/TBMD/nephritis, n (%)** | 4 (0.9) | 1 (0.6) | 3 (0) | 0 (0) | 0 (0) |
| **Family history of AS or kidney disease, n (%)** | 7 (1.7) | 5 (3.1) | 2 (4.1) | 0 (0) | 0 (0) |
| **Urology workup for hematuria, n (%)** | 75 (18.6) | 50 (31.0) | 17 (34.7) | 4 (3.4) | 4 (5.3) |
| **Genetic Testing for Alport Syndrome, n (%)** | 8 (2.0) | 5 (3.1) | 2 (4.1) | 0 (0) | 1 (1.3) |
| **Kidney Biopsy performed, n (%)** | 12 (3.0) | 9 (5.6) | 3 (4.1) | 0 (0) | 0 (0) |
| **Taking ACEi or ARBs, n (%)** | 101 (25.1) | 49 (30.4) | 18 (36.7) | 21 (17.6) | 13 (17.3) |

This table does not use USRDS data.

**Supplemental Figure 1. KDIGO Risk Categories by *COL4A3* P/LP Variant Group and Controls**

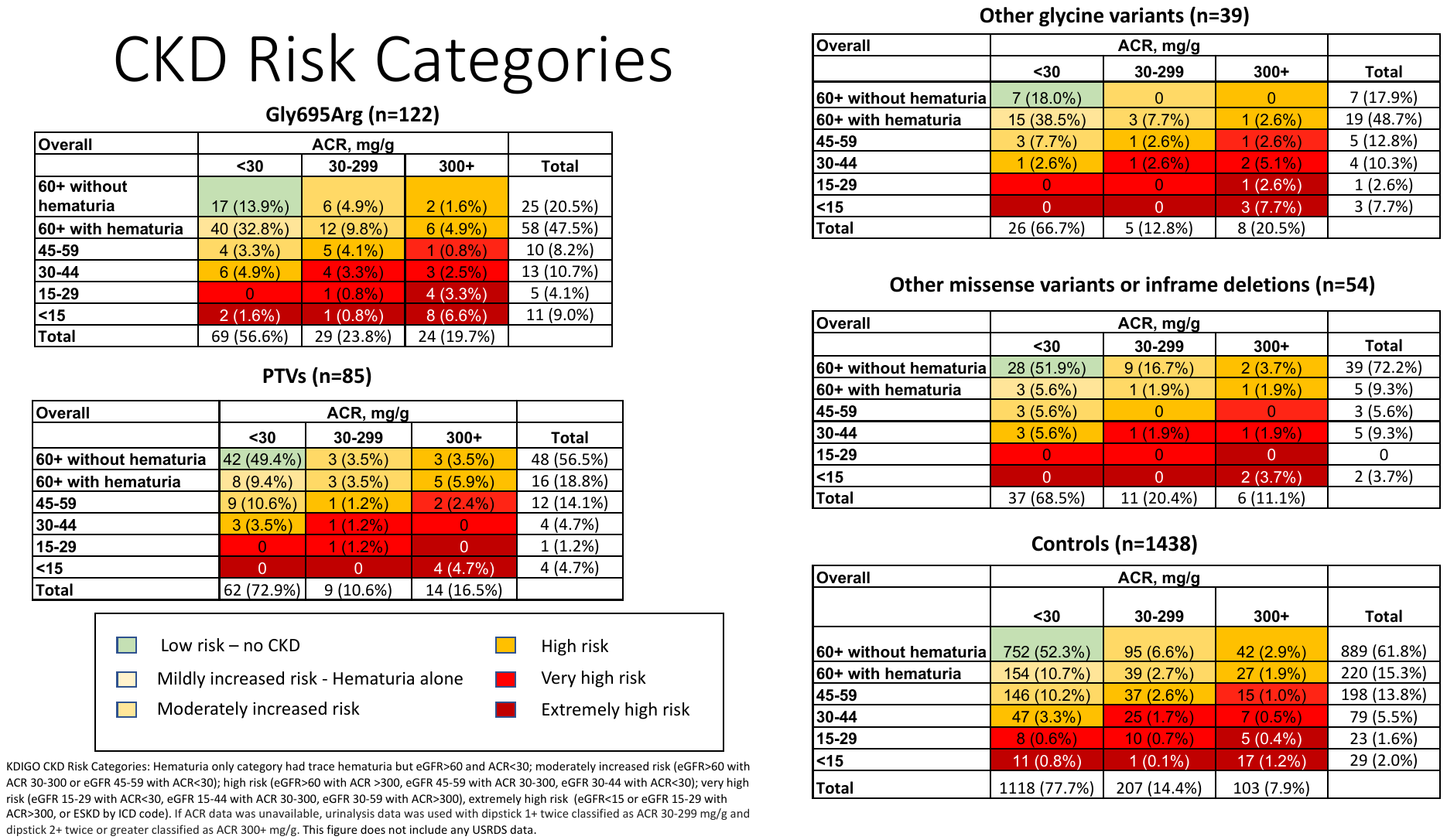

**Supplemental Figure 2. KDIGO Risk Categories in Glycine collagenous domain variants vs. controls, by age group**

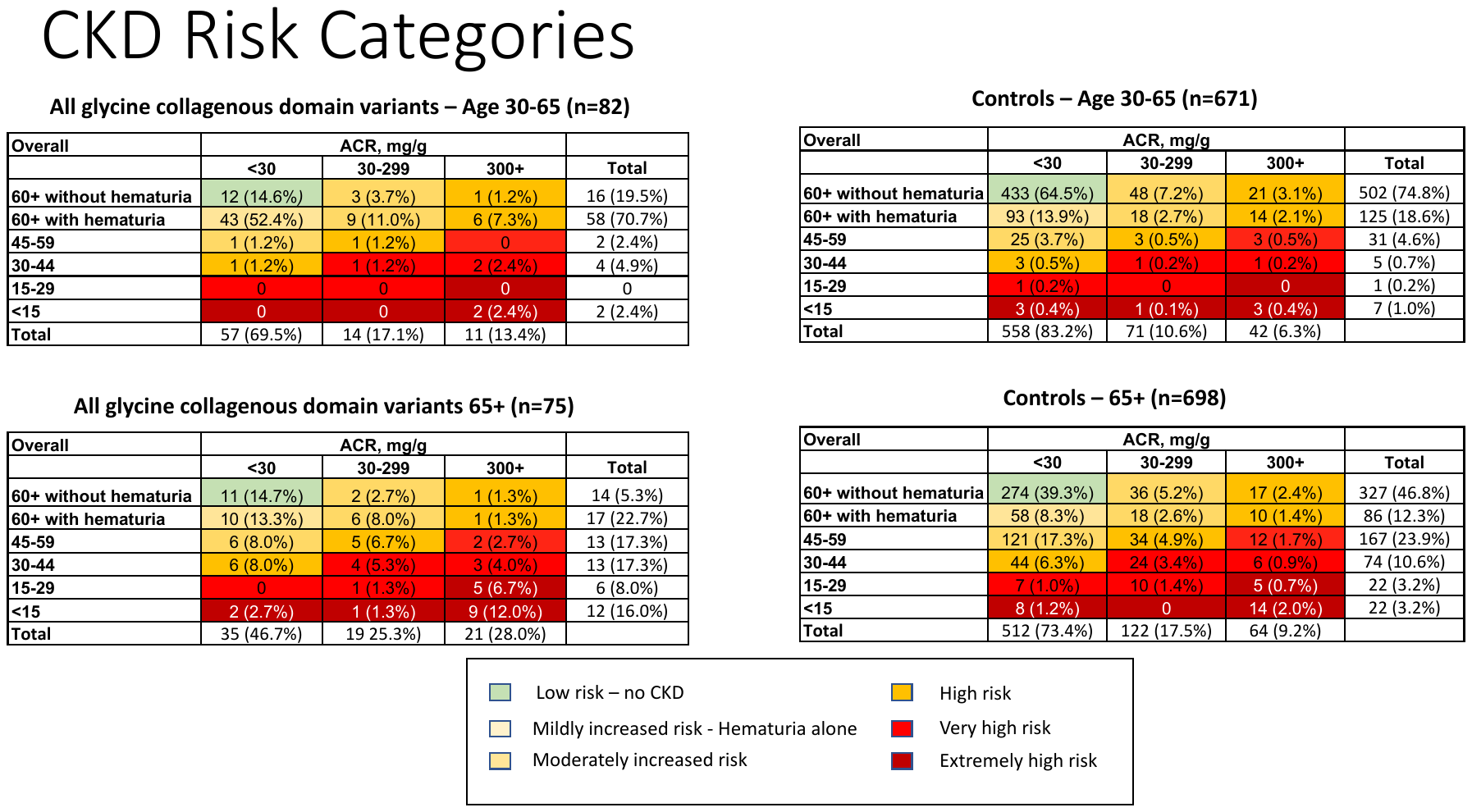
